# Implementation strategies for integrating TB treatment into community pharmacies for people with TB/HIV in Uganda using human-centered design methodology

**DOI:** 10.64898/2026.09.14.26363070

**Authors:** Jonathan Izudi, Adithya Cattamanchi, Christine Sekaggya-Wiltshire, Rachel King, Noah Kiwanuka, Amanda Sammann

## Abstract

**Background:** Community pharmacies (private retail drug shops/pharmacies) have emerged as a novel differentiated service delivery (DSD) model for delivering antiretroviral therapy (ART) to people with human immunodeficiency virus (HIV) and could support integrated tuberculosis (TB) medication refills. Using a Human-Centered Design (HCD) methodology, we developed an implementation strategy for integrating TB treatment into pharmacies targeting people with TB/HIV in Kampala, Uganda.

**Methods:** We implemented the inspiration and ideation phases of the HCD methodology. During the inspiration phase, we identified themes describing barriers and facilitators to integrating TB treatment into community pharmacies and conducted observations at community pharmacies to understand the care pathway of people with TB/HIV. We translated these qualitative findings into insight statements, design opportunities, and How Might We (HMW) questions. During the ideation phase, we conducted brainstorming and co-design workshops to generate and refine solutions, tested low-fidelity prototypes using ranked scores, and assessed the usability of high-fidelity prototypes using the System Usability Scale. Participants included people with TB/HIV, TB focal persons, HIV focal persons, Ministry of Health DSD model experts, and pharmacy healthcare providers.

**Results:** Of 26 low-fidelity prototypes, four implementation strategy components emerged: (1) raising awareness and building trust in pharmacy TB medication refills by TB and HIV focal persons, with a focus on privacy, convenience, and legitimacy; (2) standardizing TB medication refill workflows using synchronized ART/TB refill workflows, standardard operating procedures, and visual diagrams illustrating integration of TB treatment into community pharmacies; (3) strengthening the capacity of community pharmacies through certification, accreditation, and targeted TB training; and (4) strengthening monitoring and quality assurance through standard operating procedure manuals and standardized side-effect monitoring checklists.

**Conclusion:** The adapted strategy and high-fidelity prototypes will be evaluated in a pilot randomized trial assessing the effectiveness and implementation of TB treatment integration into pharmacies for people with TB/HIV in Kampala, Uganda.

## Introduction

In high tuberculosis (TB) and human immunodeficiency virus (HIV) burden settings such as Uganda, novel person-centered strategies are needed to improve TB treatment outcomes. The TB treatment success rates among people with TB/HIV in Uganda range between 54.5% and 74% according to prior studies [1–4], which is suboptimal when compared with the World Health Organization (WHO) and National TB Program (NTP) desired threshold target of ≥90%. Among several factors, missed clinic visits and long travel distances hinder the achievement of optimal treatment success and treatment adherence among people with TB/HIV. For instance, a study has shown that people with TB/HIV who travel ≥5 km to a TB clinic are less likely to achieve TB treatment success when compared with those traveling <5 km to the same clinic [5]. Another study showed that people with TB/HIV who travel ≥2 km to a TB clinic have 9% to 27% higher mortality rates when compared with those who travel less than 2 km to the same clinic [6]. Therefore, novel, context-appropriate, and person-centered strategies are critically needed to enhance TB treatment outcomes among people with TB/HIV.

Uganda implements a differentiated service delivery (DSD) model for both HIV and TB treatment in public health facilities. Evidence indicates that private retail shops (community pharmacies or pharmacies) have emerged as promising DSD models, enabling private retail pharmacies to refill antiretroviral therapy (ART) for PWH. Pharmacy-based refills have registered ≥95% ART adherence rates [7] and ≥95% viral suppression rates among PWH [8, 9]. Health care providers (HCPs) engaged in implementing pharmacy-based ART refills have reported reduced workload [7], time saved to care for PWH needing critical care [10, 11], and decongested health facilities [11]. Furthermore, because ART refills are conducted at a convenient place and time, pharmacy-based refills prevent inadvertent HIV status disclosure [12] and reduce self and community stigma associated with facility-based refills [7, 11, 13–15].

A recent review showed that pharmacy-based refills effectively deliver HIV and/or TB treatment, with documented improvements in CD4 cell counts, mean body weight, 95-100% prescription refill, 98% retention, and 99% viral suppression rates [16]. These benefits from pharmacy-based refills recorded among PWH can be extended to people with TB/HIV through community pharmacy-integrated TB/HIV medication refills. However, there are barriers and facilitators, and a critical need for a clear implementation strategy.

Our qualitative study, guided by the Consolidated Framework for Implementation Research (CFIR), showed that pharmacy-integrated TB/HIV medication refills have the potential to offer several benefits such as convenience and easy access [17], but several potential barriers, including unclear eligibility criteria for identifying and enrolling people with TB/HIV, low awareness among people with TB/HIV, and limited confidence among pharmacy providers regarding the refill strategy, among others, exist [17]. The study also reported several probable facilitators, such as the existence of a national DSD model policy, willingness of people with TB/HIV for pharmacy refills, and readiness of pharmacy providers for integrated TB/HIV medication refills, among others [17]. To leverage these facilitators and address the barriers, we proposed to adapt a contextually relevant, person-centered implementation strategy using a Human-Centered Design (HCD) methodology to inform a planned randomized trial [18]. Here, we report findings from the first two phases of the HCD methodology (inspiration and ideation), including the adapted implementation strategy for integrating TB treatment into community pharmacies for people with TB/HIV at primary health facilities in Kampala, Uganda. The third phase (implementation) will evaluate the effectiveness and implementation of this strategy in a pilot randomized trial.

## Methods and materials

### Study design and setting

This study used HCD methodology to adapt an implementation strategy for integrating TB treatment into community pharmacies for people with TB/HIV. The study was conducted across six primary healthcare facilities in Kampala, Uganda. Elsewhere [18], the study protocol is described. Each facility has an HIV clinic that is affiliated with a community pharmacy to provide ART refills to stable PWH in accordance with the Uganda Ministry of Health’s DSD model. The facilities also have TB and HIV clinics, managed by the TB focal person and HIV focal person, respectively. Both clinics provide standardized care according to the MoH national treatment guidelines. Eligible PWH who express interest in community pharmacy ART refills are identified by the focal persons in the clinics, assisted in selecting a community pharmacy of their own choice, and are thereafter linked with such pharmacies to continue with ART refills. The overall clinical care and drug supply oversight regarding ART refills is provided by the respective health facility. However, for people with TB/HIV, anti-TB refills continue through the TB clinic until treatment completion. The six facilities have been described in our earlier studies [19–23].

### Ethics statement

The Makerere University Infectious Diseases Institute Research Ethics Committee (reference number: IDI-REC-2024-98) and the Uganda National Council for Science and Technology (reference number: S4397ES) gave ethical approval. The Directorate of Public and Environmental Health, Kampala Capital City Authority, provided administrative clearance for the study (reference number: DPHE/KCCA/1301/01). All participants gave informed consent either in writing or by thumbprint after receiving comprehensive information about the study. To maintain participant privacy and confidentiality of information, no individual identifiers such as names were collected, physical documents were kept in locked cabinets, and all data were secured using a password-protected and encrypted laptop. The data remained accessible only to the research team for study purposes. Audio recordings have been transcribed verbatim and destroyed per institutional ethical guidelines. Overall, we adhered to the principles of the Declaration of Helsinki regarding the ethical conduct of research involving human participants.

### Participants and sampling approach

Participants included people with TB/HIV and healthcare providers (HCPs), namely: (1) TB focal persons, (2) HIV focal persons, (3) MoH DSD technical experts, and (4) community pharmacy HCPs. People with TB/HIV were eligible if they were aged ≥18 years, had been receiving both TB/HIV treatment for ≥2 months, and were obtaining ART refills either through community pharmacies or health facilities. The ≥2-month threshold ensured that participants had progressed beyond the intensive phase of TB treatment and had sufficient exposure to their respective ART refill models, enabling them to provide meaningful insights. Eligible people with TB/HIV were consecutively recruited from TB clinics. TB/HIV focal persons were required to have at least six months of experience providing TB care and were purposively selected. Similarly, MoH DSD technical experts with at least six months of experience in DSD programming were purposively recruited. We excluded TB/HIV focal persons who were on leave during the study period, as well as MoH experts who were unavailable. At each participating health facility, one affiliated community pharmacy serving at least 50 PWH was selected. From each pharmacy, one or two HCPs who had worked there for at least 2 months and were actively involved in ART refill services were recruited to ensure adequate experience and familiarity with the refill process. Pharmacy HCPs who were not routinely involved in ART refill services or were only temporarily covering shifts were excluded.

### Description of the HCD methodology process

We described the first two phases of the HCD methodology, namely inspiration and ideation, and adhered to the approach outlined in our published protocol [18] and a previous study [24]. These phases are informed by the Double Diamond design framework, which uses iterative processes of divergence and convergence to explore problems and develop solutions [25]. Specifically, the inspiration phase corresponded to the *Discover and Define stages*, enabling broad exploration of findings from the qualitative research (divergence) followed by synthesis into priority themes and insight statements (convergence). The ideation phase reflected the *Develop stage*, during which potential solutions were generated, refined, and prioritized through iterative expansion and narrowing of design options. Finally, the implementation phase aligned with the *Deliver stage* and focused on piloting, refining, and integrating the selected implementation strategies within real-world settings.

#### Phase 1: Inspiration phase

This phase was built on our previous qualitative study, which identified barriers and facilitators to integrating TB treatment into community pharmacies [17], complemented by observations at three community pharmacies to understand the care pathway of people with TB/HIV (journey mapping). The qualitative data were independently reviewed and synthesized by two reviewers (JI and AS). The reviewers identified themes related to the barriers and facilitators to integrating TB treatment into community pharmacies, and subsequently reviewed by a third reviewer (AC). All three reviewers (JI, AS, and AC) discussed and approved the final themes. The reviewers then generated insight statements from the themes to capture users’ perspectives, motivations, and underlying tensions, thereby ensuring specific needs of people with TB/HIV were clearly articulated. The insight statements were iteratively discussed and refined until consensus was reached among the reviewers. The insight statements informed the design opportunities and the *“How Might We”* (HMW) questions that guided the brainstorming around the identified design opportunities (actionable solutions to the identified challenges).

#### Phase 2: Ideation phase (brainstorming and prototype refinement)

The ideation phase was conducted between July 9 and 10, 2026. Stakeholders, namely people with TB/HIV, HIV and TB focal persons, MoH technical experts, and community pharmacy HCPs, participated in the 2-day co-design workshop.

During the workshop, we presented the themes, insight statements, design opportunities, HMW questions, and the low-fidelity prototypes using PowerPoint slide presentations. The presentation was followed by group brainstorming discussions involving 3-4 participants per group. The brainstorming discussions encouraged participants to challenge existing assumptions, consider multiple perspectives, and generate a wider range of contextually appropriate prototypes, including the refinement of the initial low-fidelity prototypes to better fit the local context. In this phase, we conducted two rounds of prototype testing. In the first round, we conducted low-fidelity prototype testing that involved the engagement of participants in 60-minute individual interviews with a talk-aloud session to review the prototypes. The participants thereafter privately and independently ranked the prototypes based on three criteria: (i) desirability, defined as the extent to which people wanted the prototype; (ii) feasibility, defined as the extent to which the prototype could be implemented; and (iii) viability, defined as the extent to which the prototype was sustainable and affordable. Each criterion was ranked on a 5-point Likert scale: 1 = very low, 2 = low, 3 = moderate, 4 = high, and 5 = very high (S1 File).

In the second round held on August 12, 2026, we conducted high-fidelity prototype testing using the System Usability Scale (SUS), a simple, 10-item standardized questionnaire [26] (S2 File). The participants included a subset of those involved in the low-fidelity prototype testing. Each SUS question ranked the prototype on a 1-5 scale, with the lowest score assigned to a prototype with strong disagreement and the highest score assigned to one with strong agreement. SUS was conducted until a higher level of acceptability, defined as a median score of >80.3, was reached [26]. A maximum of three iterative refinement cycles were conducted to allow systematic improvement of prototypes based on user feedback. Prototypes that achieved higher SUS were prioritized for implementation in a pilot individually randomized trial with a hybrid effectiveness-implementation design at two public primary facilities in Kampala, Uganda.

### Statistical methods: sample size and data analysis

Sample size for the inspiration phase was determined by the principle of data saturation, following the methodology established in our previous work [17]. For the subsequent ideation phase, a purposive subset of these initial participants was selected. The size and composition of this subset were determined based on expert consensus regarding participant expertise, role diversity, or thematic representation.

In the data analysis, we summarized participant demographic characteristics such as sex and level of education using frequencies and percentages and presented them in a Table. Continuous variables such as age were summarized using the mean and standard deviation when normally distributed, or the median and interquartile range (IQR) when skewed. Likert scale ratings for desirability, feasibility, and viability were summarized using the median and the interquartile range (IQR), and the overall score for each prototype was calculated using the average or mean of the three criterion median scores. The mean was then used to rank the prototypes. The analysis of the SUS data (S1 Data) involved data normalization whereby we subtracted one from each odd-numbered question (questions 1, 3, 5, 7, and 9) and subtracted five from each even-numbered question (questions 2, 4, 6, 8, and 10). The normalization process created new values that we summed/added to produce a total score and multiplied by 2.5 to yield scores that ranged from 0 to 100, with 68 as the average score [26]. We then categorized the SUS scores as ≤50, 51-68, 68-80.3, and >80.3 to suggest unacceptable, poor, good, and excellent prototypes, respectively [27]. Finally, prototypes with SUS scores ≥68 were considered usable and will be considered for testing in a planned randomized trial.

### Patient and public involvement

Patients and community members were involved in the study, particularly in a brainstorming workshop, refining and ranking prototypes based on desirability, feasibility, and viability, and evaluating the prototypes during the SUS.

### Inclusivity in global research

Supporting Information (S1 Checklist) contains information regarding ethical, cultural, and scientific considerations specific to inclusivity in global research.

## Results

### Characteristics of HCD participants

Table 1 shows the distribution of participant characteristics during the co-design workshop. We included 36 participants, equally split across the two days of the HCD workshop. The majority were from Kisenyi HC IV (n=8) and were mainly community pharmacy HCPs (n=9) or people with TB/HIV (n=9). The mean age of all participants was 36.81 years (standard deviation [SD] = 7.80). Slightly more than half were female (52.8%), nearly 45% had attained tertiary education, and approximately 92% had either formal or self-employment.

**Table 1.** Characteristics of HCD participants engaged in the HCD workshop.

| Characteristics | Level | Overall (n=36) |
| --- | --- | --- |
| Health facility | Kawaala | 6 (16.7) |
|  | Kisenyi | 8 (22.2) |
|  | Kisugu | 4 (11.1) |
|  | Kiswa | 4 (11.1) |
|  | Kitebi | 4 (11.1) |
|  | Komamboga | 4 (11.1) |
|  | MoH | 6 (16.7) |
| Type of participant | Community Pharmacy Worker | 9 (25.0) |
|  | HIV Focal Person | 6 (16.7) |
|  | MoH DSD Model Expert | 6 (16.7) |
|  | People with TB/HIV | 9 (25.0) |
|  | TB Focal Person | 6 (16.7) |
| Age group (years) | Less than 35 | 17 (47.2) |
|  | 35 and over | 19 (52.8) |
|  | mean (SD) | 36.81 (7.80) |
| Sex | Female | 19 (52.8) |
|  | Male | 17 (47.2) |
| Level of education | None | 2 (5.6) |
|  | Primary | 3 (8.3) |
|  | Secondary | 5 (13.9) |
|  | Tertiary | 16 (44.4) |
|  | University | 10 (27.8) |
| Employment status | No | 3 (8.3) |
|  | Yes | 33 (91.7) |

### Inspiration phase findings

#### Themes and illustrative quotes

From the inspiration phase, five themes emerged regarding the integration of TB treatment into community pharmacies, and they included the following:

#### Theme 1: Awareness and trust

The participants emphasized that the successful integration of TB treatment into community pharmacies would require increasing awareness and trust among both people with TB/HIV and healthcare providers.

Although the participants recognized that the refill model aligns with the MoH’s DSD model, many perceived that people with TB/HIV were unfamiliar with the model and that they might lack confidence in receiving TB treatment outside the traditional health facilities. They also highlighted that the pharmacy refill model could reduce concerns around stigma and improve acceptability. However, they emphasized the need for comprehensive patient sensitization to foster confidence in the refill model and encourage uptake.

> *It’s now the strategic direction of the Ministry of Health to integrate services… beyond just dispensing medicine.” (DSD Model Expert, Male)*.
>
> *“There are people who are wide-mouthed [meaning there are people who like rumor mongering or simply gossiping about others]… but here you move to the pharmacy and pick your drugs without anybody knowing.” (*People with TB/HIV*, Female)*.
>
> *“If the patients are not sensitized well… they might end up going back to the health facility.” (TB focal person, Female)*.

#### Theme 2: Training, workflow, and procedures

Participants highlighted the need for adequate training of community pharmacy HCPs and clearly defined implementation procedures before integrating TB treatment into community pharmacies. They emphasized that community pharmacy HCPs should receive training on TB treatment delivery and that key stakeholders, including people with TB/HIV, should be engaged early in the implementation process. They also underscored the importance of establishing clear roles and responsibilities regarding drug supply, supervision, and implementation to ensure consistent, coordinated, and high-quality service delivery.

> *“They should involve us before starting it [TB treatment integration into community pharmacies]. We can help explain to others.” (*Person with TB/HIV*, Male)*.

#### Theme 3: Accessibility and counseling

Participants viewed community pharmacies as a convenient and accessible platform for TB treatment because of their extended working hours, shorter waiting times, and greater privacy compared with public health facilities. These characteristics were perceived as likely to improve convenience and reduce stigma associated with seeking TB care. However, concerns were raised regarding the capacity of community pharmacy HCPs to safely store TB medications and provide appropriate counseling.

> “*Most community pharmacies open at 8:30 am… some work 24 hours, unlike health facilities.” (TB focal person, Male)*.
>
> *“In the health facility, there are many people [high patient volume]… but in the community pharmacy, you pick your drugs and leave [because there are fewer patients].” (*Person with TB/HIV*, Female)*.

#### Theme 4: Incentives for pharmacy HCPs

Participants reported that community pharmacy HCPs were generally willing to implement TB treatment integration into community pharmacies, although they emphasized that sustained engagement would likely require appropriate incentives. In this regard, both financial and non-financial incentives were reported as important motivating factors for providers, including recognizing providers’ additional responsibilities and ensuring that TB services remain a priority within pharmacy settings. The participants suggested adapting existing incentive models used for HIV DSD models to support implementation of pharmacy-based TB treatment.

> *“Like for ART [Antiretroviral therapy] integration [of TB treatment] into community pharmacies, the organization gives us 2,000 Uganda shillings for every patient… I think the same can be done with TB treatment.” (Community pharmacy HCP, Male)*.

#### Theme 5: Protocol workflow for TB medication dispensing

Participants highlighted the importance of developing standardized implementation protocols to guide community pharmacy-based integrated HIV and TB drug refills. They raised concerns around drug procurement, inventory management, accountability, supervision, and reporting. They recommended a need for clear operational procedures supported by electronic systems to facilitate accountability for drugs, strengthen communication between pharmacies and health facilities, and ensure safe and consistent delivery of integrated refills through community pharmacies.

> *“We need a clear implementation plan. Who supplies the drugs? Who trains us? Who supervises us?” (Community Pharmacy HCP, Male)*.
>
> *“How do we ensure that these pharmacies will account for the medicines properly? We need an electronic system…” (DSD Model Expert, Male)*.

### Ideation phase findings

#### Insight statements, design opportunities, and How Might We (HMW) questions

Based on emergent themes in Table 1, we generated five insight statements that informed the design opportunities (Table 2). The insight statements present the synthesis of participants’ underlying needs and contextual challenges. The *“How Might We (HMW)”* questions guided the brainstorming of the design opportunities, including: (1) increasing awareness and trust in pharmacy-based TB refills, (2) streamline workflows and clarifying procedures for TB service delivery in community pharmacies, (3) improving accessibility and patient counselling, (4) developing sustainable incentive mechanisms for community pharmacy healthcare providers, and (5) establishing clear protocols for TB medication dispensing and accountability. Five design opportunities corresponding to each insight statement were developed along with 26 low-fidelity prototypes.

**Table 2.**
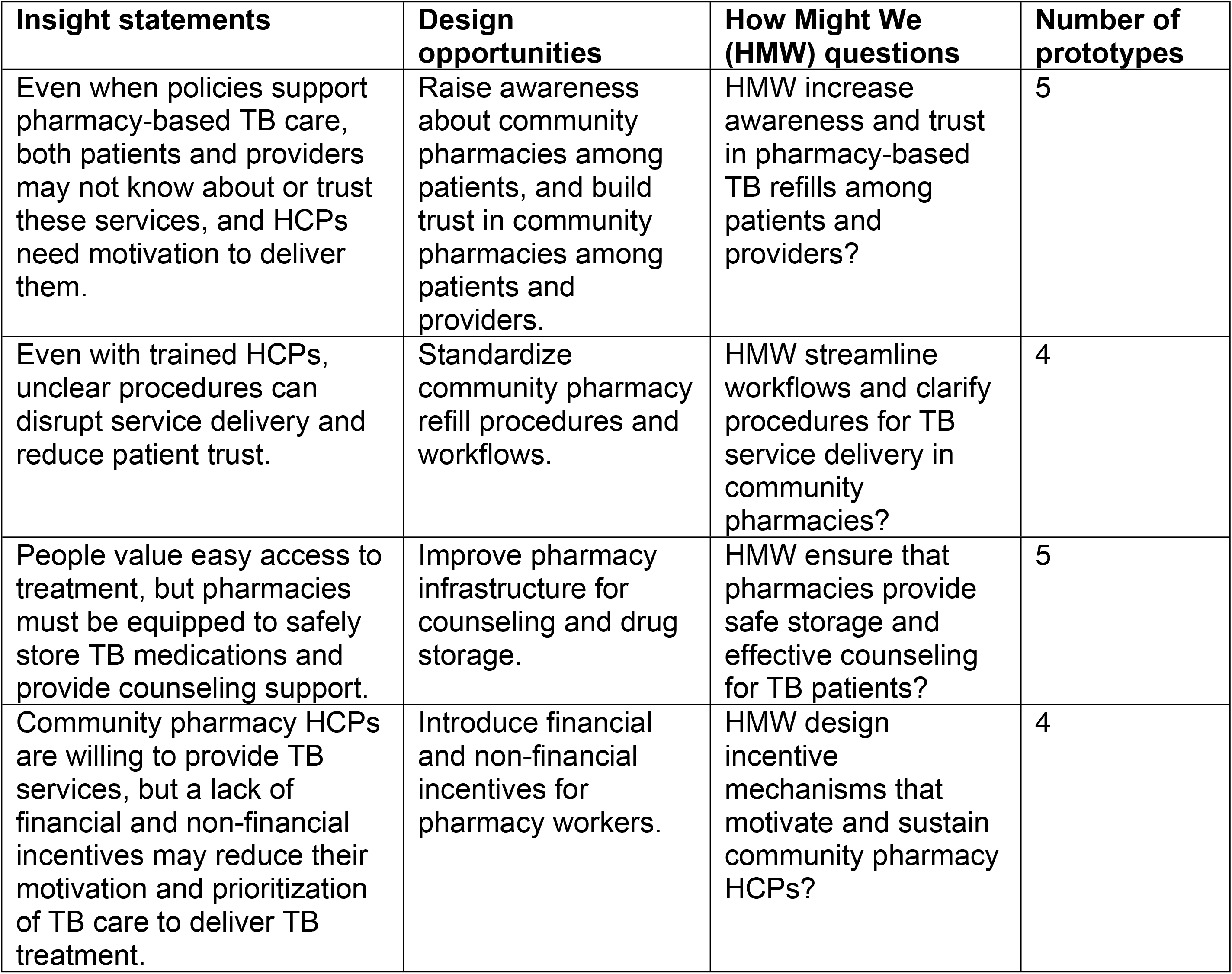
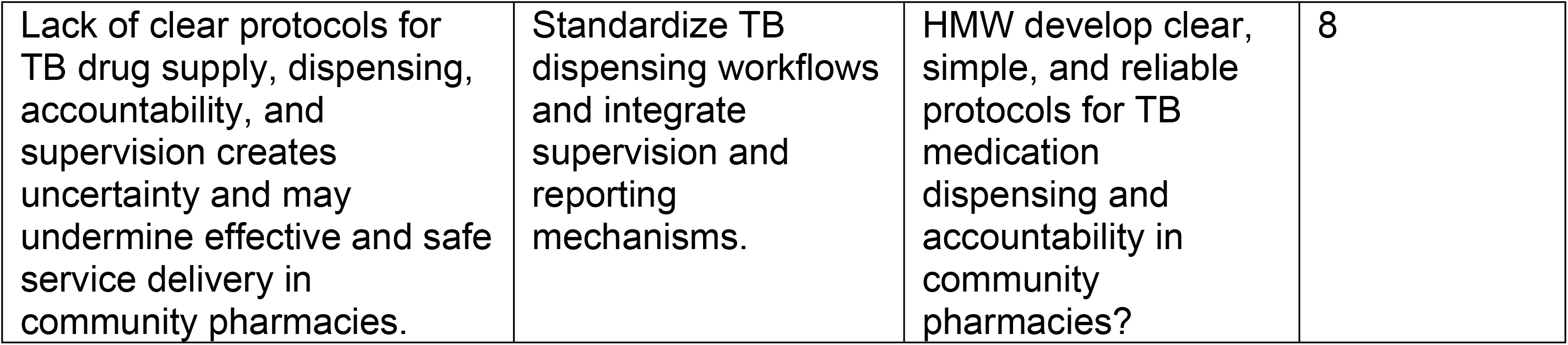
Insight statements, design opportunities, and How Might We (HMW) questions.

#### Low-fidelity prototype testing

The five design opportunities (DO) and their initial low-fidelity prototypes were reviewed and refined. Table 3 presents the selected refined prototypes and their rankings, based on their strengths, contextual considerations, and implementation limitations. Of the 26 prototypes, nine (34.6%) were selected, with none selected from the third design opportunity.

**Table 3:**
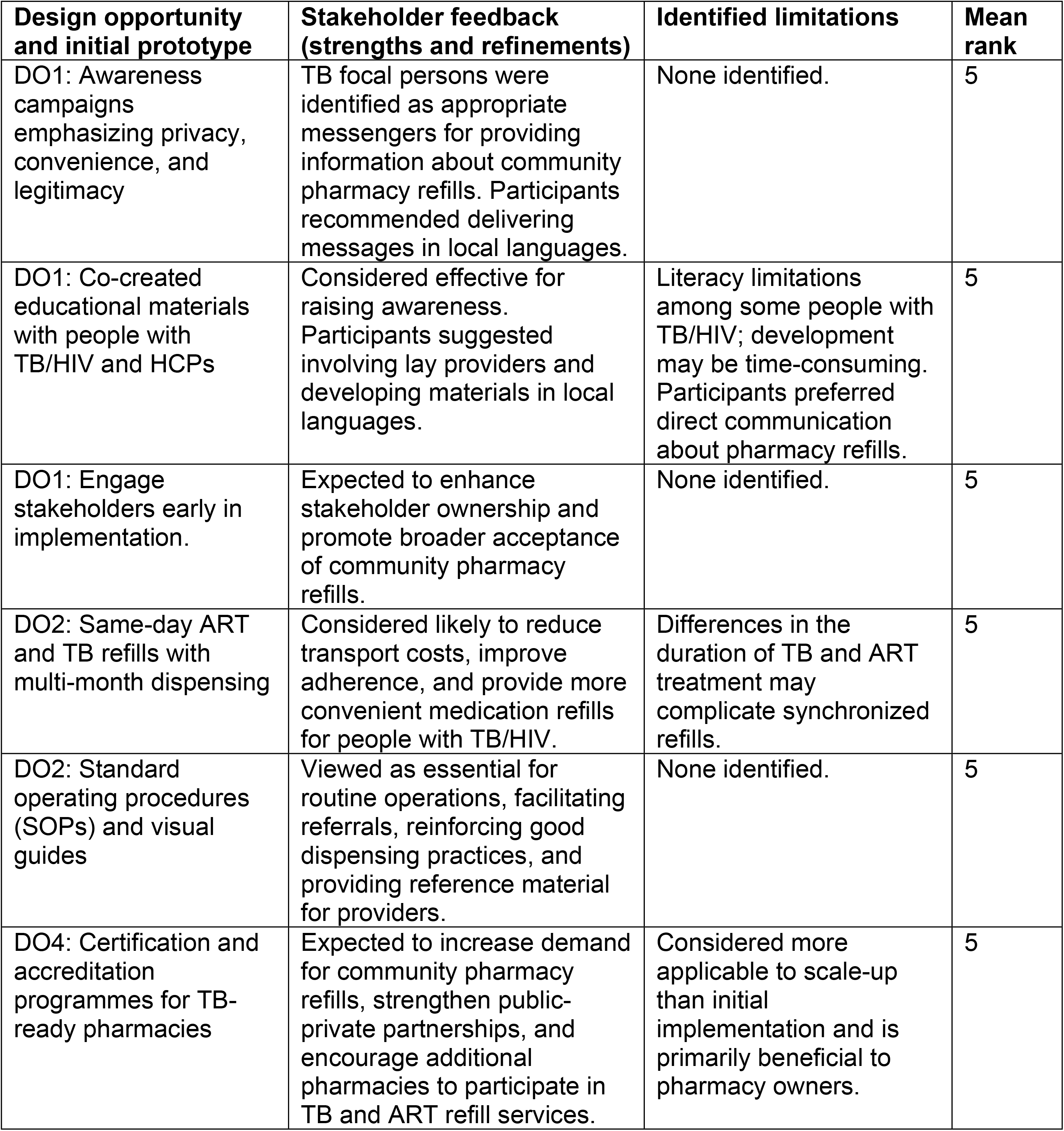
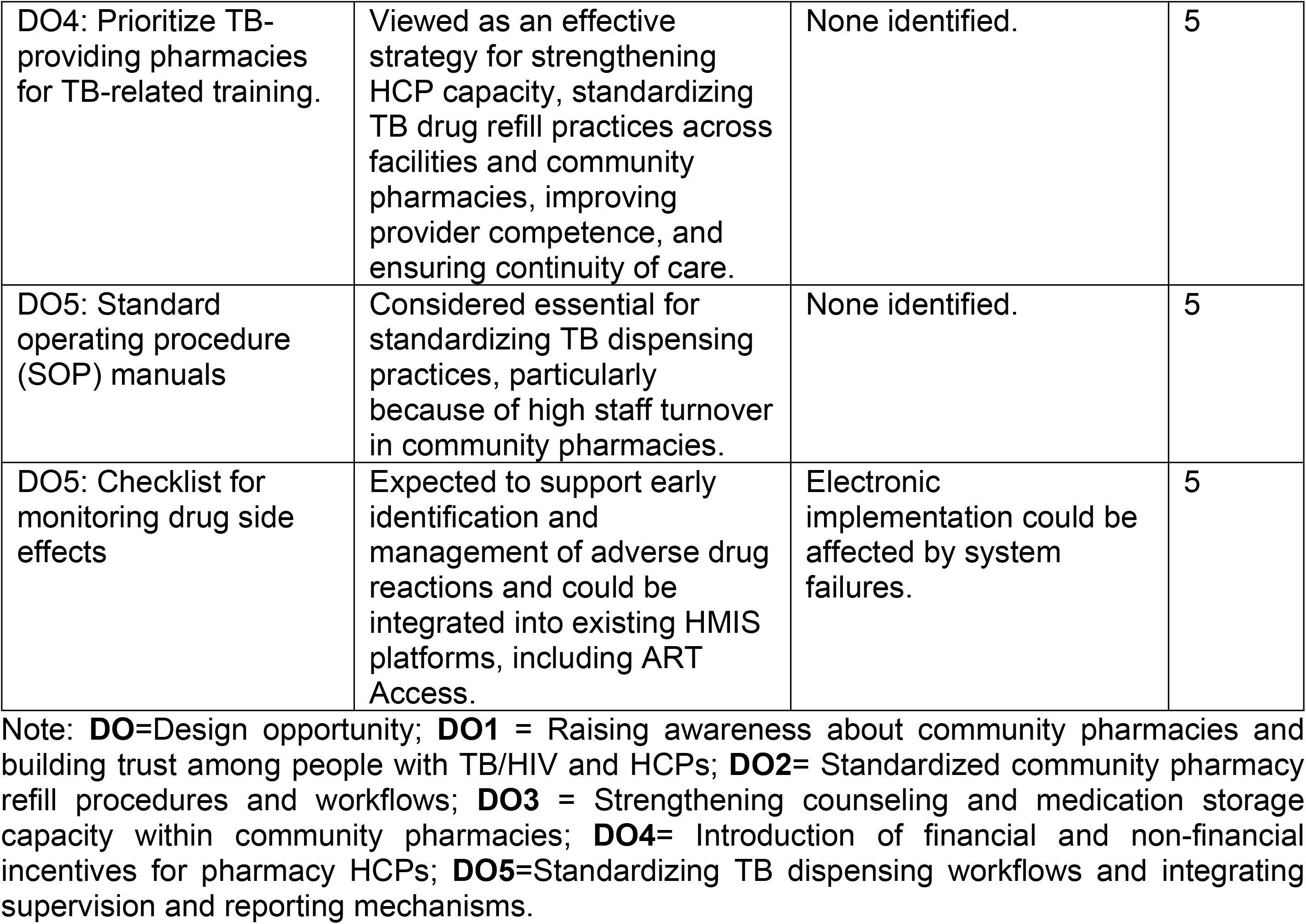
Design opportunity matrix of selected refined prototypes and their rankings.

| **Design opportunity and initial prototype** | **Stakeholder feedback (strengths and refinements)** | **Identified limitations** | **Mean rank** |
| --- | --- | --- | --- |
| DO1: Awareness campaigns emphasizing privacy, convenience, and legitimacy | TB focal persons were identified as appropriate messengers for providing information about community pharmacy refills. Participants recommended delivering messages in local languages. | None identified. | 5 |
| DO1: Co-created educational materials with people with TB/HIV and HCPs | Considered effective for raising awareness. Participants suggested involving lay providers and developing materials in local languages. | Literacy limitations among some people with TB/HIV; development may be time-consuming. Participants preferred direct communication about pharmacy refills. | 5 |
| DO1: Engage stakeholders early in implementation. | Expected to enhance stakeholder ownership and promote broader acceptance of community pharmacy refills. | None identified. | 5 |
| DO2: Same-day ART and TB refills with multi-month dispensing | Considered likely to reduce transport costs, improve adherence, and provide more convenient medication refills for people with TB/HIV. | Differences in the duration of TB and ART treatment may complicate synchronized refills. | 5 |
| DO2: Standard operating procedures (SOPs) and visual guides | Viewed as essential for routine operations, facilitating referrals, reinforcing good dispensing practices, and providing reference material for providers. | None identified. | 5 |
| DO4: Certification and accreditation programmes for TB-ready pharmacies | Expected to increase demand for community pharmacy refills, strengthen public-private partnerships, and encourage additional pharmacies to participate in TB and ART refill services. | Considered more applicable to scale-up than initial implementation and is primarily beneficial to pharmacy owners. | 5 |
| DO4: Prioritize TB-providing pharmacies for TB-related training. | Viewed as an effective strategy for strengthening HCP capacity, standardizing TB drug refill practices across facilities and community pharmacies, improving provider competence, and ensuring continuity of care. | None identified. | 5 |
| DO5: Standard operating procedure (SOP) manuals | Considered essential for standardizing TB dispensing practices, particularly because of high staff turnover in community pharmacies. | None identified. | 5 |
| DO5: Checklist for monitoring drug side effects | Expected to support early identification and management of adverse drug reactions and could be integrated into existing HMIS platforms, including ART Access. | Electronic implementation could be affected by system failures. | 5 |

Note: **DO**=Design opportunity; **DO1** = Raising awareness about community pharmacies and building trust among people with TB/HIV and HCPs; **DO2**= Standardized community pharmacy refill procedures and workflows; **DO3** = Strengthening counseling and medication storage capacity within community pharmacies; **DO4**= Introduction of financial and non-financial incentives for pharmacy HCPs; **DO5**=Standardizing TB dispensing workflows and integrating supervision and reporting mechanisms.

Selected prototypes from the first design opportunity included: i) awareness campaigns emphasizing privacy, convenience, and legitimacy; ii) educational materials co-developed with people with TB/HIV and HCPs and communicated directly to people with TB/HIV to address literacy barriers; and iii) early stakeholder engagement in designing TB treatment integration in community pharmacies. From the second design opportunity, i) same-day ART and TB medication refills with multi-month dispensing was selected because of its potential to reduce transport costs, improve adherence, and increase convenience for people with TB/HIV; and ii) standard operating procedures (SOPs) and visual guides demonstrating routine operations, provider guidance, and referral processes were selected.

From the fourth design opportunity, two prototypes were selected: certification and accreditation of TB-ready pharmacies and prioritization of participating pharmacies for TB-related training. Participants provided two reasons for the selection. First, these approaches were considered sustainable options for strengthening pharmacy capacity, standardizing TB drug refills, and reinforcing public-private partnerships. Second, public recognition of participating pharmacies was considered likely to increase stigma by making people with TB/HIV more identifiable. Lastly, under the fifth design opportunity, SOP manuals and checklists for monitoring drug side effects were prioritized for further development because they were perceived as practical and sustainable and as capable of improving the quality and consistency of pharmacy-based TB drug refills. Participants emphasized that SOPs could standardize dispensing despite frequent staff turnover, while side-effect monitoring checklists could facilitate early identification and management of adverse events and be integrated into existing health information systems. S3 File presents the excluded prototypes.

### High-fidelity prototype testing using SUS

#### Characteristics of participants engaged in the SUS

Table 4 shows the characteristics of the 15 participants who engaged in the SUS evaluation of the prototypes. Participants comprised TB focal persons, HIV focal persons, community pharmacy HCPs, people with TB/HIV, and MoH DSD model experts. The median age was 35.0 years (IQR 31.5-39.5), with 53.3% of participants being female. Two-thirds (66.7%) had university-level education, and the median duration of work experience was 6.0 years (IQR 5.0-9.3, n=13).

**Table 4:** Characteristics of participants engaged in the SUS.

| Variable | Level | Overall (n=15) |
| --- | --- | --- |
| Participant | Community Pharmacy HCP | 3 (20.0) |
|  | HIV focal person | 4 (26.7) |
|  | MoH DSD Model Experts | 2 (13.3) |
|  | People with TB/HIV | 2 (13.3) |
|  | TB focal person | 4 (26.7) |
| Health facility | Kawaala | 3 (20.0) |
|  | Kisenyi | 2 (13.3) |
|  | Kisugu | 1 (6.7) |
|  | Kiswa | 2 (13.3) |
|  | Kitebi | 2 (13.3) |
|  | Komamboga | 3 (20.0) |
|  | MoH | 2 (13.3) |
| Age categories (years) | Less than or equal to 35 | 7 (46.7) |
|  | Over 35 | 8 (53.3) |
|  | Median (IQR) | 35.0 (31.5-39.5) |
| Sex | Female | 8 (53.3) |
|  | Male | 7 (46.7) |
| Highest level of education | Tertiary | 5 (33.3) |
|  | University | 10 (66.7) |
| Work experience (years) | Less than 5 | 6 (40.0) |
|  | 5 and more | 6 (40.0) |
|  | Not reported | 3 (20.0) |
|  | Median (IQR) | 6.0 (5.0-9.3) |

#### SUS findings regarding high-fidelity prototypes

Based on the rankings of the low-fidelity prototypes, four high-fidelity prototypes were developed and evaluated by the participants using the SUS (Table 5). The prototypes included: (1) TB drug side-effect monitoring checklist (S4 File); (2) visual workflow for TB medication dispensing at the community pharmacy (S1 Fig). S1 Fig: Visual workflow for TB medication dispensing at the community pharmacy; (3) stakeholder co-designed integrated same-day ART and TB medication refill workflow (S2 Fig). S2 Fig: Stakeholder co-designed integrated same-day ART and TB medication refill workflow; and (4) community pharmacy SOP for medication refills (S5 File).

**Table 5:** SUS findings regarding high-fidelity prototypes.

| Prototype | Total | Mean (SD) | Median (IQR) |
| --- | --- | --- | --- |
| Side effect monitoring checklist | 15 | 95.23 (6.84) | 97.5 (93.75-100.0) |
| Visual workflow for TB medication dispensing at the community pharmacy | 15 | 85.23 (9.18) | 82.5 (77.5-90.0) |
| Stakeholder co-designed integrated same-day ART and TB medication refill workflow | 15 | 86.14 (10.33) | 85.0 (76.25-95.0) |
| Community pharmacy SOP for medication refills. | 15 | 90.23 (4.93) | 90.0 (87.5-93.75) |

The usability testing demonstrated high usability across all four co-designed prototypes. The side effect monitoring checklist had the highest mean SUS score (95.23, SD 6.84; median 97.5, IQR 93.75-100.0), followed by the standard operating procedure (90.23, SD 4.93; median 90.0, IQR 87.5-93.75), co-designed diagram (86.14, SD 10.33; median 85.0, IQR 76.25-95.0), and community pharmacy TB and ART workflow (85.23, SD 9.18; median 82.5, IQR 77.5-90.0). All prototypes scored well above the conventional SUS benchmark of 68, indicating excellent perceived usability.

#### Implementation strategy for integrating TB treatment into community pharmacies

Based on the refinement and prioritization of low-fidelity prototypes and subsequent SUS evaluation, we retained eight prototypes and synthesized them into a four-component implementation strategy to support the integration of TB medication refills into community pharmacies (Table 6). The implementation strategy components included: (1) raising awareness about community pharmacies and building trust in community pharmacy TB medication refills by TB and HIV focal persons, with a focus on privacy, convenience, and legitimacy of the refill model; (2) standardizing medication refill workflows using same ART and TB refill workflows with multi-month dispensing, standard operating procedures (SOPs), and visual diagrams illustrating the integration of TB treatment into community pharmacies; (3) strengthening the capacity of community pharmacies through certification, accreditation, and targeted TB training by the Uganda MoH NTLP; and (4) strengthening monitoring and quality assurance through SOP manuals and standardized side-effect monitoring checklist.

**Table 6:** Implementation strategy for integrating TB treatment into community pharmacies.

| Implementation strategy component | Rationale for the implementation strategy component | Key components developed through co-design workshop | Responsible person/ entity |
| --- | --- | --- | --- |
| **Component 1.**<br>Raise awareness about community pharmacies and build trust in community pharmacy TB medication refills | To increase awareness about community pharmacies and improve acceptability, including building trust among people with TB/HIV and HCPs at both facilities and | TB and HIV focal persons raise awareness about pharmacy refills, emphasizing privacy, convenience, and legitimacy using appropriate languages. Stakeholder engagement | TB and HIV focal persons |
|  | community pharmacies regarding pharmacy refills. | throughout the implementation of community pharmacy-based refills. |  |
| **Component 2.**<br>Standardize TB and ART medication refill workflows | To seamlessly improve the efficiency, consistency, and integration of TB medication refills within community pharmacies. | Same-day ART and TB medication refills with multi-month dispensing; SOPs and visual diagrams to guide dispensing and referrals. | TB and HIV focal persons |
| **Component 3.**<br>Strengthen community pharmacy capacity through accreditation, certification, and targeted TB training | To build the capacity and readiness of community pharmacies to provide standardized TB medication refills. | Certification and accreditation of TB-ready pharmacies completed and should continue during scale-up by the MoH and National TB Program (NTP). Participating community pharmacies receive orientation on integrated TB and ART refills and are prioritized for TB-related training. | MoH and NTP |
| **Component 4.**<br>Strengthen monitoring and quality assurance using a standardized TB drug side effect monitoring checklist | To improve the quality and safety of TB and ART medication refills through standardized monitoring and ensure quality care. | SOP manuals for pharmacy HCPs and a checklist for monitoring TB drug side effects during medication refills. | Community pharmacy HCPs. |

## Discussion

Using the HCD methodology, we adapted a person-centred strategy to integrate TB treatment into community pharmacies for people with TB/HIV in Kampala, Uganda. We generated four contextually grounded implementation strategy components that stakeholders considered feasible and acceptable for HIV and TB management, aligning with the goal of HCD to provide an innovative and person-centred approach to developing implementation strategies [28]. Adoption of the implementation strategy involved interactive and participatory decision-making among key stakeholders, which may positively influence its acceptability, fidelity, and feasibility [29–33]. The strategy may also contribute to the anticipated benefits of integrating TB treatment into community pharmacies, including reduced self- and community-level stigma associated with TB and HIV[7, 11, 13–15], reduced inadvertent disclosure of TB and HIV status [12], reduced workload [7], more time to care for people with TB/HIV requiring additional support [10, 11], decongestion of health facilities [11], and greater convenience [34].

Several lessons emerged from this HCD work, alongside a few constraints. First, despite its value in tailoring interventions to user needs, HCD remains underutilized in TB implementation research. Second, effective implementation requires early identification of suitable individuals to participate in its various phases. Third, clearly explaining the study aims and rationale for HCD was critical. Establishing this understanding early minimized ambiguity and facilitated focused stakeholder discussions. Fourth, meaningful HCD workshops require highly skilled facilitators who can manage the fluid design process and steer participant engagement towards actionable outputs. Lastly, involving institutional experts from project inception is vital for securing long-term programmatic buy-in and sustainability. For example, MoH DSD model experts participated in all HCD phases and provided guidance on the feasibility and practicality of several prototypes to support long-term implementation fidelity and sustainability.

One major operational challenge was stakeholder mobilization. Competing priorities and demanding schedules could significantly affect attendance and participation. We mitigated this challenge through a multi-channel communication strategy, including introductory emails sent one week before the HCD workshop, digital reminders 2-3 days before the event, direct telephone calls, and physical facility visits conducted 1-2 days before the workshop.

### Implications of the study findings for future research and policy

We will evaluate the effectiveness and implementation outcomes of the adapted implementation strategy components in a pilot, individually randomized trial with a type 2 hybrid effectiveness-implementation design. The effectiveness outcomes will include HIV viral load suppression to demonstrate optimal response to ART and TB treatment success to measure optimal TB program performance under pharmacy refill conditions. The implementation outcomes will include feasibility of implementing integrated TB and ART medication refills, including acceptability and fidelity. The evidence generated from the pilot randomized trial will inform the scale-up of community pharmacy TB medication refills across Uganda through a cluster randomized trial.

### Study strengths and limitations

Our study has several strengths. All relevant stakeholders involved in TB management and community pharmacy-based service delivery participated, enabling the identification of context-specific needs, preferences, and challenges that might otherwise have been overlooked by researcher-driven approaches. Their inclusion also provided perspectives from frontline service delivery, program implementation, and the lived experiences of people with TB/HIV, which may improve the relevance, feasibility, adoption, sustainability, and acceptability of the implementation strategy. Furthermore, iterative prototyping and real-time feedback supported the development of strategies that were responsive and adaptable to the context. Lastly, perspectives from marginalized and underrepresented groups were equitably represented in developing and adopting the implementation strategies.

Despite these strengths, the study has limitations. First, HCD requires skilled facilitators and multiple rounds of stakeholder engagement, which can be resource-and time-intensive. We addressed these challenges by engaging facilitators experienced in HCD methodology and using structured, well-coordinated stakeholder engagement processes. Second, the prototypes developed may be context-specific and may not be generalizable to other settings. We addressed this limitation through iterative prototype refinement and high-fidelity prototype testing using the SUS. Third, incomplete stakeholder representation and selection bias may not have been eliminated despite involving participants at multiple levels of TB care delivery. Finally, HCD may generate user-centered solutions that do not fully address structural or health system barriers. We mitigated this limitation by integrating HCD with the Consolidated Framework for Implementation Research (CFIR), a determinant implementation science framework, during the qualitative phase. This approach ensured that the implementation strategies were informed by both user and health system perspectives.

### Conclusion and recommendation

We adapted a person-centered implementation strategy for integrating TB treatment into community pharmacies for people with TB/HIV in Kampala, Uganda, using an HCD methodology. The process resulted in four contextually grounded implementation strategy components: 1) raising awareness about community pharmacy refills and building trust in community pharmacy refills by TB and HIV focal persons with focus on privacy, convenience, and legitimacy; (2) standardizing TB medication refill workflows using synchronized ART and TB refill workflows with multi-month dispensing, SOPs, and visual diagrams illustrating the integration of TB treatment into community pharmacies; (3) strengthening community pharmacy HCP capacity through certification, accreditation, and targeted TB training by the MoH and NTP; and (4) strengthening monitoring and quality assurance through SOP manuals and standardized TB medication side-effect monitoring checklists. These strategies will guide the implementation of TB treatment integration into community pharmacies in a planned pilot individually randomized trial targeting people with TB/HIV at two primary health facilities in Kampala, Uganda.

## Data Availability

All relevant data are within the manuscript and its Supporting Information files

## Supporting information

S1 Checklist: Inclusivity in global research.

S1 File: System Usability Scale for high-fidelity prototype testing.

S1 Data. System Usability Data.

S2 File: System Usability Scale for high-fidelity prototype testing.

S3 File: Excluded prototypes.

S4 File: TB drug side-effect monitoring checklist.

S5 File: Community pharmacy SOP for medication refills.

S1 Fig: Visual workflow for TB medication dispensing at the community pharmacy.

S2 Fig: Stakeholder co-designed integrated same-day ART and TB medication refill workflow.

---

**S1 Fig.**
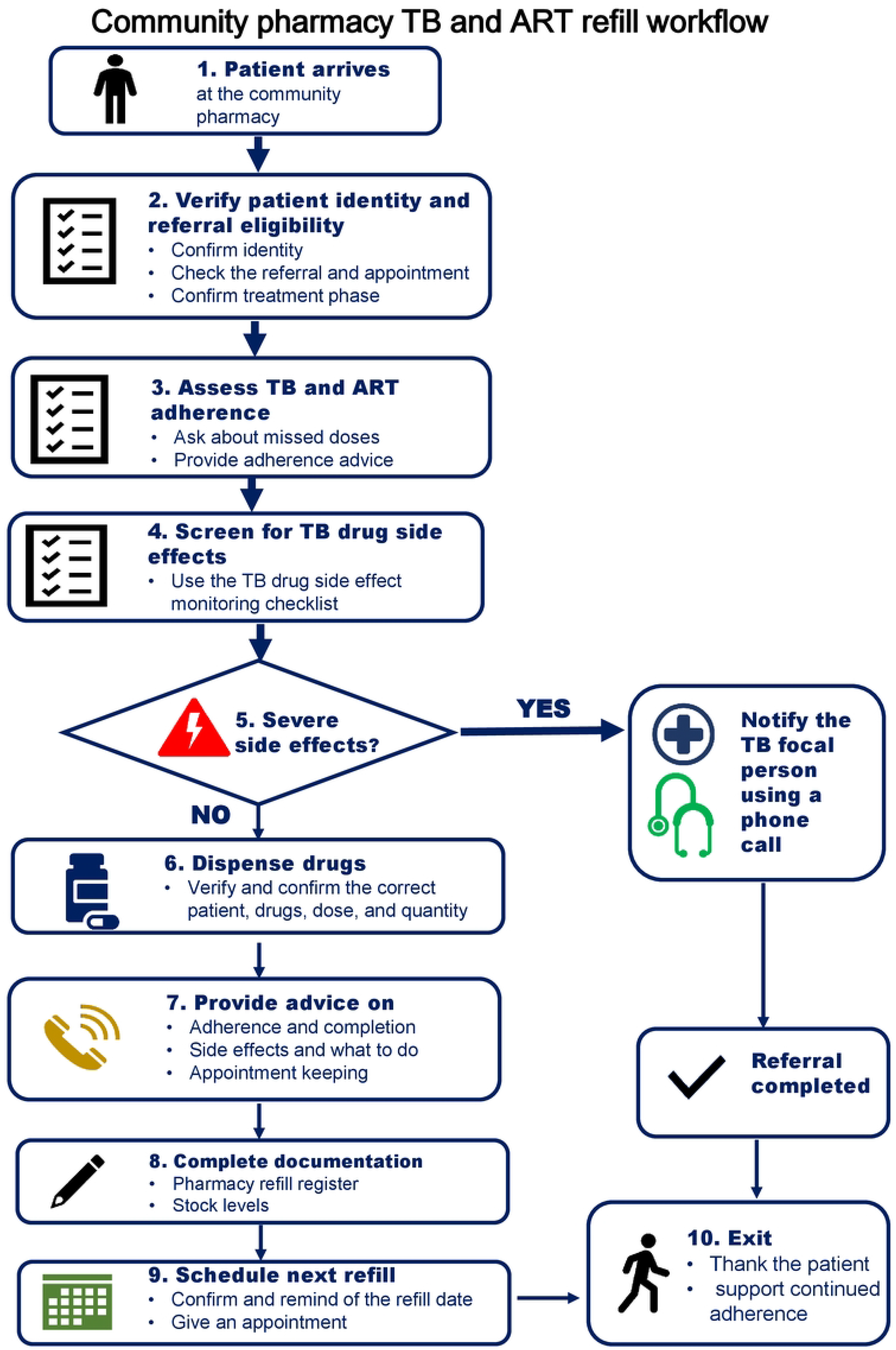
Workflow for TB medication dispensing at community pharmacies. The figure shows the workflow for TB medication dispensing at community pharmacies, from arrival of people with TB/HIV to completion of the pharmacy visit. Upon arrival, records are verified, TB and ART adherence are assessed, and TB drug side effects are evaluated. For those without side effects, TB and ART medications are dispensed, counseling and advice are provided, documentation is completed, and the next refill date is scheduled. For those experiencing severe TB drug side effects, the TB focal person is notified, and referral to the health facility is arranged by telephone.

**S2 Fig.**
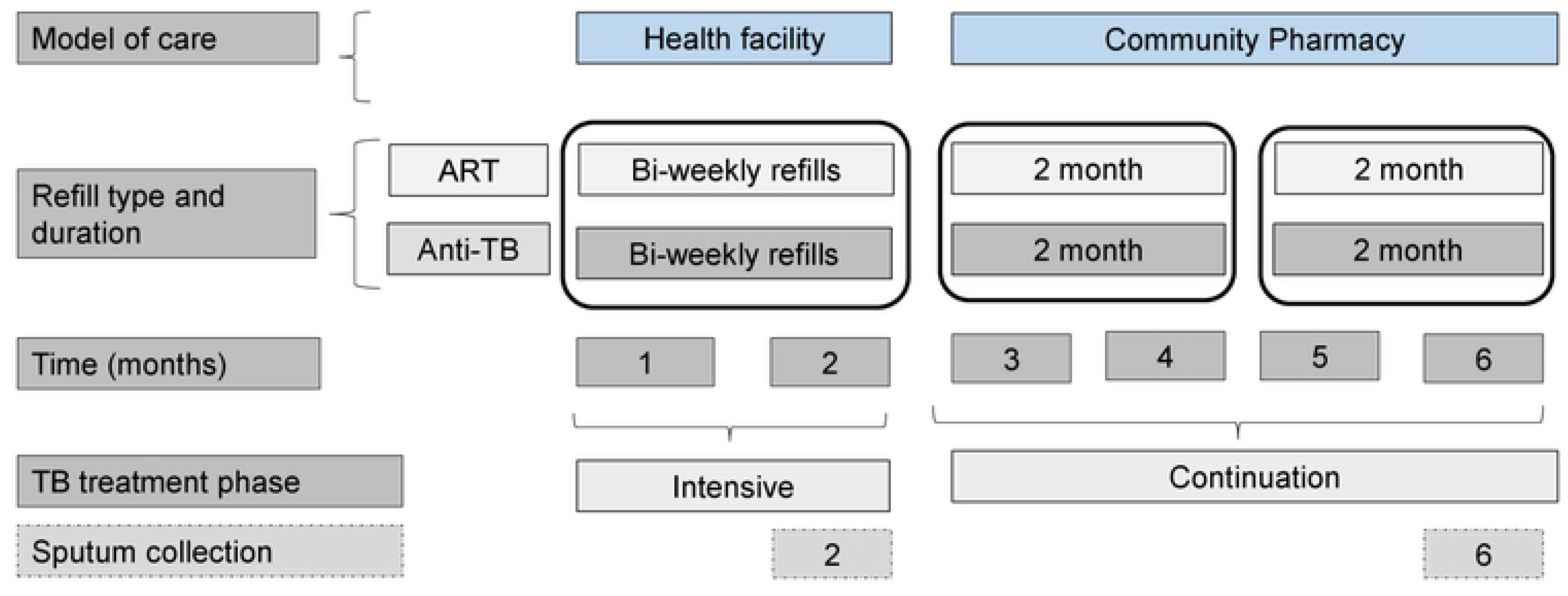
Stakeholder co-designed integrated same-day ART and TB medication refill workflow. The figure presents the stakeholder co-designed workflow for integrated same-day ART and TB medication refills. During the first 2 months, people with TB/HIV are assessed for stability at each TB clinic visit and receive standardized TB counseling and health education, adherence assessment and support, and sputum smear follow-up examinations if they have bacteriologically confirmed pulmonary TB. During this period, both ART and TB medications are refilled biweekly. At month 2, eligible people with TB/HIV are randomized to either community pharmacy-based care or treatment as usual (routine care). Before randomization, all participants with bacteriologically confirmed pulmonary TB undergo a sputum smear examination at the end of month 2. In the community pharmacy arm, ART and TB medications are subsequently refilled twice, with each refill providing a 2-month supply. At the end of month 6, participants undergo a sputum smear examination through a laboratory-only visit, following referral from the pharmacy to the health facility for ART refill, after which they return to community pharmacy-based refills. In the treatment-as-usual arm, TB medications are refilled monthly between months 3 and 6, while ART medications are refilled every 2-3 months, depending on the multi-month dispensing strategy implemented at the facility. Participants in the treatment-as-usual arm undergo sputum smear examinations at the end of months 5 and 6 according to routine care.

